# Practice of Hyperglycaemia Control in Intensive Care Units of the Military Hospital, Sudan – Needs of a Protocol

**DOI:** 10.1101/2020.08.17.20176453

**Authors:** Ghada Omer Hamad Abd El-Raheem, Mudawi Mohammed Ahmed Abdallah, Mounkaila Noma

**Affiliations:** Intensive Care Unit, Military Hospital, Khartoum, Sudan. & University of Medical Sciences and Technology UMST, High Diploma in Research Methodology and Biostatistics, Khartoum, Sudan. P.O. Box 12810, Mecca street, Khartoum, Sudan. /; Intensive Care Unit, Military Hospital, Medical Manager of Critical Care Department. Military Hospital, Omdurman, Khartoum, Sudan.; University of Medical Sciences and Technology, P.O. Box 12810, Mecca street, Khartoum, Sudan. E-mail.

**Keywords:** Local protocol, Hyperglycaemia control, Intensive Care Unit, critically ill, Sudan

## Abstract

Hyperglycaemia is a major risk factor in critically ill patients as it leads to adverse outcomes and mortality in diabetic and non-diabetic patients. The target blood glucose remained controversial; this study aimed to contribute in assessing the practice of hyperglycaemia control in intensive care units of Khartoum Military Hospital. Furthermore, it proposed a protocol for hyperglycaemia control based on findings. A hospital-based cross-sectional study assessed the awareness and practice towards hyperglycaemia management in a sample of 83 healthcare staff selected through stratified random sampling technique. In addition, 55 patients were enrolled, through quota sampling, after excluding those with diabetic ketoacidosis, hyperosmolar-hyperglycaemic state and patients < 18 years. A self-administrated questionnaire enabled to collect data from healthcare staff, patients data were extracted from medical records. SPSS 23 was used to analyse the collected data. Chi-square and ANOVA tests assessed the association among variables. All statistical tests were considered statistically significant when *p* < 0.05. The training on hyperglycaemia control differed statistically (*p* = 0.017) among healthcare staff. The target glycaemic level (140-180 mg/dl) was knew by 11.1% of the study participants. Neither the knowledge nor the practice of hyperglycaemia control methods differed among staff (*p*> 0.05). The use of sliding scale was 79.3% across the ICUs with a statistically significant difference (*p* = 0.002). 31.5% of patients had received glycaemic control based on different methods and 11.8% were in the targeted blood glucose level. Sliding scale was the prevalent method used by doctors (71.4%) and nurses (81.6%). A patient benefited from insulin infusion method, which achieved the NICE-SUGAR target. The poor knowledge and lack of awareness towards hyperglycaemia monitoring led to inappropriate implementation of glycaemia control methods across the Military Hospital ICUs. Sustained training programs on hyperglycaemia control to ICU staff and the availability of a protocol on glycaemia control are highly required.

## Introduction

Hyperglycaemia is a major risk factor affecting critically ill patients leading to adverse outcomes and a high mortality in diabetic and non-diabetic patients [1–4]. Stressful situations, as acute illness and surgery in particular neurosurgery, elevate the levels of stress hormones and increase hepatic glucose production, lipolysis and insulin resistance [5–7]. The stress cascade increases by 7-8 folds in patients undergoing surgery [8, 9] leading to more than three folds increase in post-surgical complications and by six folds for mortality [7]. The target blood glucose (BG) had been controversial. Leuven 1 study was the first landmark clinical trial that revealed the benefits of reduced morbidity and mortality related to intensive insulin therapy (IIT) in surgical critically ill patients [3, 10]. However, the second Leuven trial with a higher hypoglycaemia rate, pointed out that in medical intensive care unit (ICU) patients, there was a no statistically significant difference in mortality rate between tight blood glucose group (80-110 mg/dl) and control group [2, 3, 10]. In 2009, the practice changed following the publication of the Normoglycaemia in Intensive Care Evaluation (NICE) and Survival Using Glucose Algorithm Regulation (SUGAR) trial [11]. It revealed that the mortality and the hypoglycaemia increased in the intensive insulin therapy group compared to the conventional group. Furthermore, a subgroup analysis indicated a no difference in outcomes between medical, surgical, diabetic, non-diabetic and septic patients [2, 3, 7]. With respect to these results, conventional target of < 180 mg/dl was acceptable for most ICU patients [4] and adopted by various professional organizations [2, 3, 12, 13], except the American College of Physicians (ACP) which proposed a higher target of BG (< 200 mg/dl) [14]. These controversial benefits from intensive insulin therapy should not shadow the reduction of complications and length of hospitalization in hepatobiliary-pancreatic surgical patients [15]. Intravenous insulin through infusion pump is the method applied for ICU patients [11, 13]. The concomitant use of sub-cutaneous insulin glargine remains more efficacious than insulin infusion alone, in particular in patients with coronary artery bypass graft [16,17].

For policy development, it is crucial to assess the current practice by surveying both healthcare staff and patients to identify barriers and facilitators through a gap analysis to establish the best practice [18]. Regarding hyperglycaemia control policy, the major safety issue remained hypoglycaemia, especially in ICU patients, as the usual symptoms might not be noticed [19]. Hypoglycaemia defines as blood glucose level < 70 mg/dl and severe life threatening when it is < 40 mg/dl [11, 20, 21].

Protocols were developed as written instructions to prevent the fluctuation in BG due to changes of interventions as administering steroids, vasopressors or parenteral quinine or due to changes in nutrition support [22]. As hyperglycaemia is more prevalent in patients receiving parenteral nutrition [9], BG levels did not differ among eating and non-per oral (NPO) patients [23]. The protocols had differences in their target BG, monitoring frequencies, infusion rates and use of boluses [24]. Hence, they must be customized to suit local resources, staff competency [25] and the needs of patients [25–27]. Examples of these protocols are Portland l, Washington University, and Yale University protocols. Yale Protocol had more difficult calculations than the other protocols [28], however, its hypoglycaemia rate was lower than Leuven protocol [22]. The Nottingham University Hospitals (NUH) protocol adopted BG target levels, which were consistent with the NICE-SUGAR target [13].

Alternative approaches to written policies are computerized protocols such as glucommanders [28], star protocol [4] and space glucose control (SGC) system [29]. Although, they reduced the nursing workload and had lower hypoglycaemia rates [21], they had not changed the general practice [27].

This study assessed the practice of healthcare staff on hyperglycaemia control in intensive care units of Khartoum Military Hospital and proposed a protocol for hyperglycaemia control from lessons learnt.

## Materials and Methods

A hospital-based cross-sectional study assessed the awareness, and practice of healthcare staff towards hyperglycaemia management and the burden of hyperglycaemia control based medical records of critically ill patients in the intensive care units of the Military Hospital of Khartoum State, Sudan. The Military Hospital is a complex of seven specialized hospitals totalizing 722 beds and 8 ICUs. A multistage sampling technique was used. At first level, five ICUs were systematically included in the study after excluding the neonatal, the maternity and the medical ICUs the last being under reconstruction. At second level, a stratified random sampling technique enabled to select 83 health professionals (doctors and nurses) proportionally to the size of each ICU after excluding the administrative staff. Regarding the patients included in the study, a quota of 12 patients was fixed to randomly recruit participants from each of the five ICUs. This led to an estimated sample of 60 patients. Fifty-five patients were enrolled in the study after excluding those with either diabetic ketoacidosis (DKA) or hyperosmolar hyperglycaemic state (HHS) and patients < 18 years. Data were collected through a standardized questionnaire comprising two parts. Part one was a self-administrated questionnaire filled by the healthcare staff working in ICUs to collect their sociodemographic characteristics, their number of years of working experience, their knowledge and practice on hyperglycaemia control methods and levels as well as the management of hyperglycaemia. Part two extracted data from the medical records of ICU patients hospitalized at the time of the data collection. The characteristics of the patients: age, gender, status (medical or surgical), type of hyperglycaemia (diabetes type 1, 2 or non-diabetic), associated comorbidities, methods of blood glucose measurement and levels were recorded. The statistical package for social sciences (SPSS version 23) was used to describe and analyse the data. Statistical analysis performed were chi-square tests and analysis of variance (ANOVA) to determine association among variables. All tests were considered statistically significant when *p* < 0.05.

## Results

### Characteristics of the Healthcare Staff and their training on hyperglycaemia control

The majority (74.1%, 60/81) of the participants were nurses and the remaining 25.9% (21/81) were doctors. 77.8% (63/81) of the participants were aged 25-30 years with no statistical association (*p* = 0.05) between the age of the participants and their occupation as indicated by table 1. The years of working experience of the participants ranged between 0.1 year and 12 years with median of 1 year; while, working years in intensive care unit ranged from 0.01 years to 8 years with a median of working years of 0.5 years. 66.7% (14/21) of the doctors received training on hyperglycaemia control and 36.7% (22/60) of the nurses were trained with a statistically significant difference (5.67, *p* = 0.017) between the status and being trained on hyperglycaemia.

**Table 1:** Characteristics of the healthcare staff and training on glycaemic control (n=81)

| Characteristics | Status of the staff |  |  |  |  |  | Likelihood ratio | p-value |
| --- | --- | --- | --- | --- | --- | --- | --- | --- |
|  | Doctor | % | Nurse | % | Total | % |  |  |
| **Age:** |  |  |  |  |  |  |  |  |
| 25-30 years | 13 | 20.6 | 50 | 79.4 | 63 | 77.8 | 3.835 | 0.05 |
| > 30 years | 8 | 44.4 | 10 | 55.6 | 18 | 22.2 |  |  |
| **Total (%)** | **21** | **25.9** | **60** | **74.1** | **81** | **100.0** |  |  |
| **Gender:** |  |  |  |  |  |  |  |  |
| Female | 19 | 28.8 | 47 | 71.2 | 66 | 81.5 | 1.697 | 0.193 |
| Male | 2 | 13.3 | 13 | 86.7 | 15 | 18.5 |  |  |
| **Total (%)** | **21** | **25.9** | **60** | **74.1** | **81** | **100.0** |  |  |
| **ICU working experience:** |  |  |  |  |  |  |  |  |
| <1 year | 11 | 20.4 | 43 | 79.6 | 54 | 66.7 | 4.849 | 0.089 |
| 1-3 years | 9 | 45.0 | 11 | 55.0 | 20 | 24.7 |  |  |
| >3 years | 1 | 14.3 | 6 | 85.7 | 7 | 8.6 |  |  |
| **Total (%)** | **21** | **25.9** | **60** | **74.1** | **81** | **100.0** |  |  |
| **Training about Glycaemic control:** |  |  |  |  |  |  |  |  |
| Trained | 14 | 38.9 | 22 | 61.1 | 36 | 44.4 | 5.67* | 0.017 |
| Untrained | 7 | 15.6 | 38 | 84.4 | 45 | 55.6 |  |  |
| **Total (%)** | **21** |  | **60** |  | **81** | **100.0** |  |  |

\*chi-square test

### Awareness of healthcare staff towards the target blood glucose level

The 81 healthcare staff were asked if they knew the target blood glucose (BG) level, 88.9% (72/81) replied yes, 27.8% (20/72) of them were doctors and 72.2% (52/72) were nurses. They were 11.1% (9/81) who did not know, 11.1% (1/9) were doctors and 88.9% (8/9) were nurses. There was a no statistically significant association (Likelihood ratio = 1.349, *p* = 0.245) between the awareness about target BG level and the staff status. However, when prompted to provide the exact level of the target blood glucose, they were 11.1% (8/72) who provided the correct level (140-180 mg/dl) and 88.9% (64/72) reported incorrect levels. Of the eight participants who reported the correct level, 62.5% (5/8) were doctors and 37.5% (3/8) were nurses. A statistically significant difference (Fisher’s Exact Test, *p* = 0.033) was found between the reported level of blood glucose and the status of the healthcare staff.

### Awareness of healthcare staff about Basal-Bolus and Insulin infusion methods

Regarding the hyperglycaemia control methods, Basal-Bolus and Insulin Infusion, there was no statistical significant association between having training or not on hyperglycaemia control methods and health profession with a *p-value* of respectively 0.591 and 0.371 (table 2).

**Table 2:**
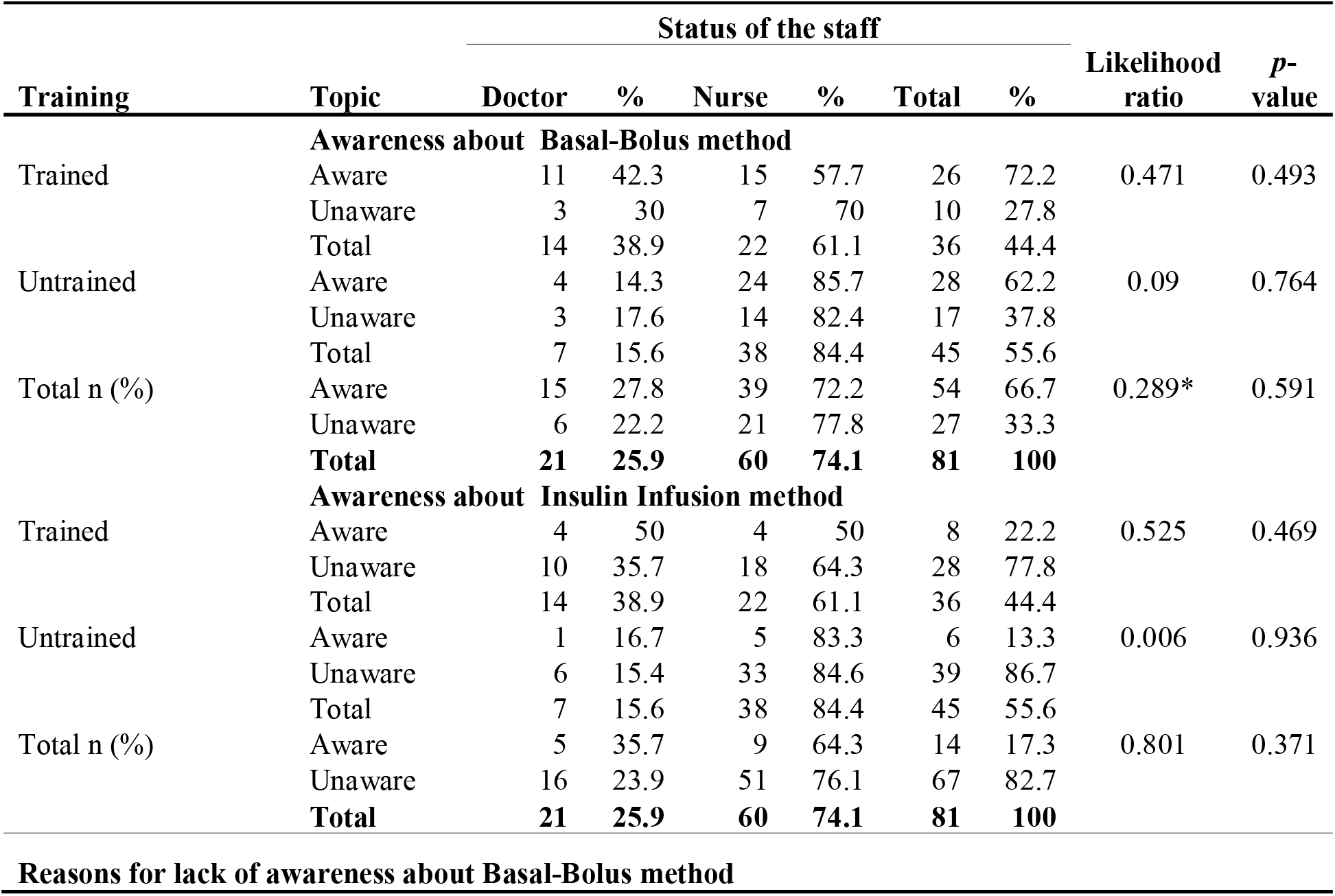
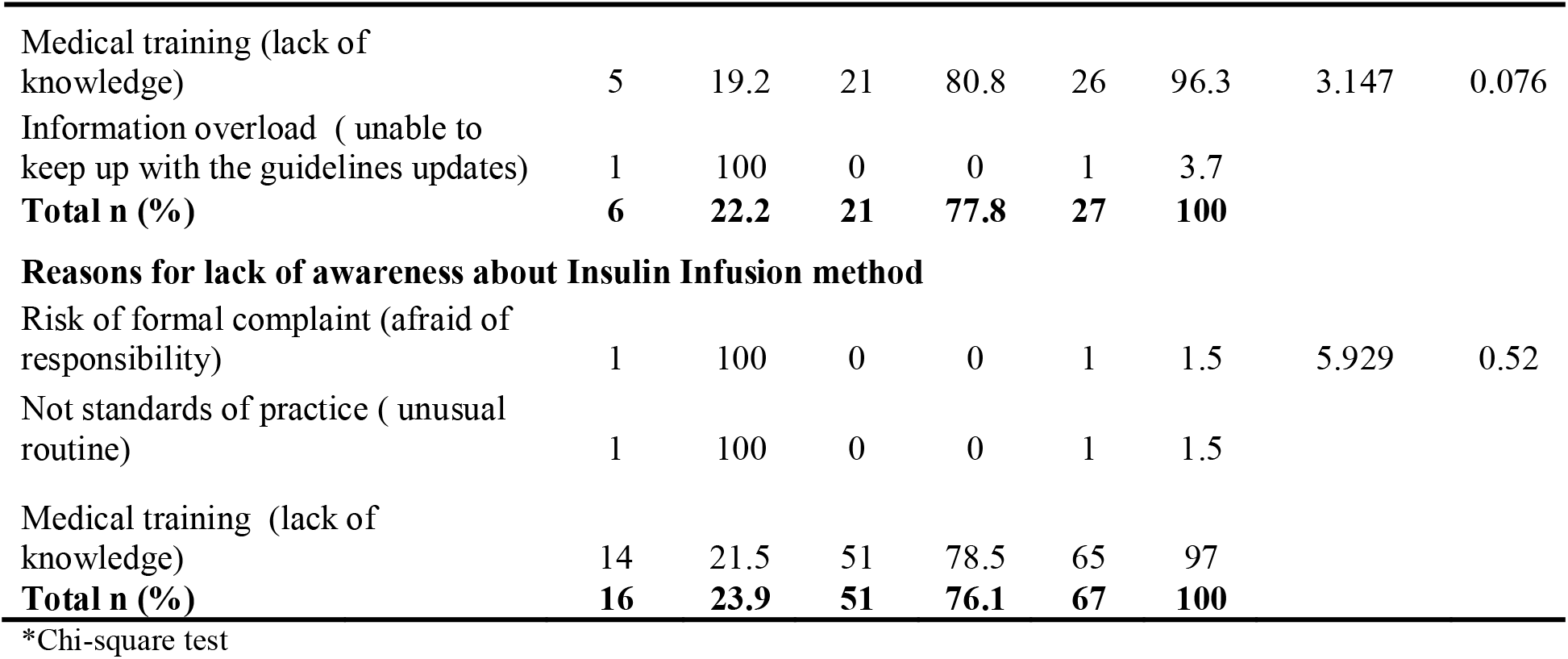
Awareness of health care staff towards hyperglycaemia control methods and the reasons for lack of awareness (n=81)

Of the 27 staff members (6 doctors and 21 nurses) who did not know about *Basal-Bolus method*, the main reason was the lack of knowledge reported by 96.3% (26/27) of the staff. There was no statistically significant difference (Likelihood ratio = 3.147, *p* = 0.076) between the reasons of lack of awareness and the profession of healthcare staff (table 2). Regarding *Insulin Infusion method*, of the 67 staff members (16 doctors and 51 nurses) who did not know about it, the main reason was also the lack of knowledge reported by 97 % (65/67) of the staff. There was a no statistically significant difference (Likelihood ratio = 5.929, *p* = 0.52) between the reasons for lack of awareness and the status of staff pas revealed by table 2.

### Practice of healthcare staff towards glycaemic control Practice towards blood glucose monitoring frequency

The practice of staff towards blood glucose (BG) measurement was assessed as either more frequently (< 6 hourly) or less frequently (≥ 6 hourly).

Regarding doctors, 47.6% (10/21) measured BG more frequently, among them, 80% (8/10) were trained on hyperglycaemia control and 20% (2/10) were not. The remaining 52.4% (11/21) measured BG level less frequently, 54.5% (6/11) were trained and 45.5% (5/11) were untrained. There was no statistically significant association (Likelihood ratio = 1.567, *p* = 0.211) between the training status of doctors and their practice toward BG monitoring frequency.

With regard to nurses, 35.0% (21/60) measured BG more frequently, among them, 42.9% (9/21) received training on hyperglycaemia control and 57.1% (12/21) did not. The remaining 65.0% (39/60) measured BG level less frequently, 33.3% (13/39) were trained and 66.7% (26/39) were untrained. A non-statistically significant association (χ^2^ = 0.533, *p* = 0.465) was found between the training status of nurses and their practice toward BG monitoring frequency.

In the overall, a no statistically significant association was found (χ^2^ = 2.197, *p* = 0.138) between the training of staff and their practice towards BG monitoring frequency.

### Management of diabetic ketoacidosis

In the overall, the appropriate management of diabetic ketoacidosis (DKA), consisting of overlapping the I.V and S.C insulin was performed by 29.6% (24/81) of the participants. The remaining 70.4% (57/81) either they stopped the I.V insulin then start the S.C insulin (56.8%, 46/81) or they did not know what to do (13.6%, 11/81); with a statistically significant difference between doctors and nurses as revealed by table 3.

**Table 3:** Management of diabetic ketoacidosis (DKA) by the participants according to their status of training on glycaemia control (n=81)

| **DKA Management** | **Doctor** | **%** | **Nurse** | **%** | **Health Staff** | **%** | **Likelihood ratio** | **P-value** |
| --- | --- | --- | --- | --- | --- | --- | --- | --- |
| *Trained* |  |  |  |  |  |  |  |  |
| Stop the I.V insulin then start the S.C insulin | 4 | 21.1 | 15 | 78.9 | 19 | 52.8 | 6.627 | 0.036 |
| Overlap the I.V and S.C | 9 | 56.3 | 7 | 43.8 | 16 | 44.4 |  |  |
| Do not know | 1 | 100 | 0 | 0 | 1 | 2.8 |  |  |
| **Total** | **14** | **38.9** | **22** | **61** | **36** | **100** |  |  |
| *Untrained* |  |  |  |  |  |  |  |  |
| Stop the I.V insulin then start the S.C insulin | 4 | 14.8 | 23 | 85.2 | 27 | 60 | 5.663 | 0.059 |
| Overlap the I.V and S.C | 3 | 37.5 | 5 | 62.5 | 8 | 17.8 |  |  |
| Do not know | 0 | 0 | 10 | 100 | 10 | 22.2 |  |  |
| **Total** | **7** | **15.6** | **38** | **84** | **45** | **100** |  |  |
| *Trained and untrained* |  |  |  |  |  |  |  |  |
| Stop the I.V insulin then start the S.C insulin | 8 | 17.4 | 38 | 82.6 | 46 | 56.8 | 10.229 | 0.006 |
| Overlap the I.V and S.C | 12 | 50 | 12 | 50 | 24 | 29.6 |  |  |
| Do not know | 1 | 9.1 | 10 | 90.9 | 11 | 13.6 |  |  |
| **Total** | **21** | **25.9** | **60** | **74.1** | **81** | **100** |  |  |

### Practice of healthcare staff towards glycaemia measurement and control methods used in the different Intensive care units

Across the three types of intensive care units, *HbA1c measurement* was requested by 69.5% (57/82) of the health staff and the remaining 30.5% (25/82) did not. In cardiac care unit (CCU), the request was from all (8/8) the staff; while in mixed and surgical ICUs it was respectively from 71.9% (46/64) and 30% (3/10) of the staff. There was a statistically significant association (Likelihood ratio = 12.584, *p* = 0.002) between ICU type and the request of HbA1c measurement.

*Sliding scale method* was used by 79.3% (65/82) of all the ICU staff, 90% (9/10) of the surgical ICU staff, 84.4% (54/64) of the mixed and 25% (2/8) of the cardiac ICUs. In the overall, across these three types of ICU, sliding scale method was used by 79.3% (65/82) of the staff and they were 20.7% (17/82) who used other methods. These other methods used by the remaining 17 staff members were Basal-Bolus method (82.3%, 14/17), mixed insulin method (11.8%, 2/17) and insulin infusion (5.9%, 1/17). There was a statistically significant association (Likelihood ratio = 12.728, *p* = 0.002) between the use of sliding scale method and the type of the ICU.

### Hyperglycaemia control methods used by health care professionals

Figure 1 revealed the distribution of staff by hyperglycaemia method used. The sliding scale was the prevalent method used by both doctors and nurses with respectively 71.4% (15/21) and 81.6% (49/60). The other control methods (basal bolus, insulin Infusion and mixed insulin) were used by respectively 28.6% (6/21) and 18.3% (11/60) of the doctors and nurses. A no statistically significant difference (Likelihood ratio = 0.938, *p* = 0.333) was found between hyperglycaemia control method used and the position of healthcare staff.

**Figure 1:**
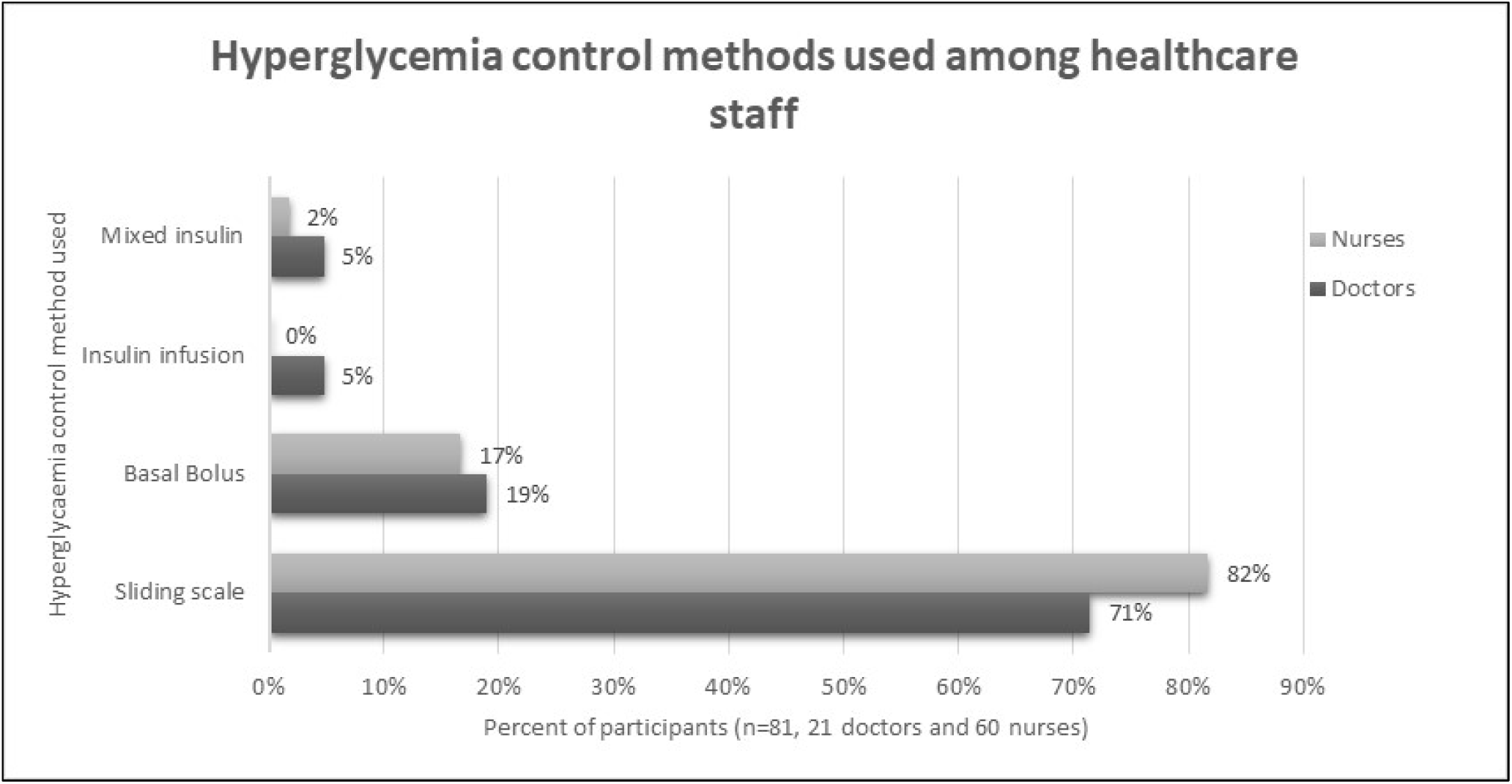
Hyperglycemia control methods used among healthcare professionals

When asked to provide reasons for using either sliding scale or other hyperglycaemia control methods, 80.5% (66/82) of the staff reported that it was the usual practice, 18.3% (15/82) because it was instructed by the first level supervisor, information overload was provided as the main reason by a participant (1.2%, 1/82).

Of the sixty-six participants who reported standard practice as their main reason, the majority 97.0% (64/66) used sliding scale and the remaining 3.0% (2/66) used other glycaemia control methods. A statistically significant association (Likelihood = 65.778, *p* = 0.000) was found between the reasons of using a particular control method and glycaemia control methods.

The staff were requested to provide their opinion on the control methods used, their opinion was recoded as satisfied (yes), not satisfied (no) and no opinion (do not know). More than half (53.7%, 44/82) were not satisfied with the control method they were using, they were 41.5% (34/82) who were satisfied and 4.9% (4/82) could not provide a justification. A no statistical significant association (Likelihood = 2.018, *p* = 0.365) was found between the control method used and the satisfaction towards the method used.

### Number of infusion pumps per ICU

The number of infusion pumps available per ICU patient ranged from 0 to 6 with a statistically significant mean infusion pumps of 2.95 ± 1.33 (*p* = 0.001) across the five ICUs (3 mixed, 1 cardiac and 1 surgical). This average varied across the ICUs, the lowest mean was recorded in the surgical unit with a mean of 1.70 pumps±0.82 [range: 0-3]. The cardiac unit had an average of 2.25 pumps±1.39 [range: 1 – 5]. The mixed ICUs were more equipped with an average of 2.86 pumps±0.86 [range: 2-5] in Room-B, 3.15 pumps±1.03 [1–6] in CCR1 and 3.57 pumps±1.6 [1–6] in CCR2.

### Characteristics of ICU patients

Of the Fifty-five patients selected across the ICUs of the Military Hospital, 50.9% were males and 49.1% were females. They were between 19 and 95 years with a median age of 63.5 years. More than half (58.2%, 32/55) were under enteral feeding. Their glycaemia status indicated that 72.8% (40/55) were non-diabetic; 23.6% (13/55) and 3.6% (2/55) were respectively type 2 and type 1 diabetic patients. Table 4 displayed the other characteristics of the patients.

**Table 4:** Characteristics of the patients hospitalized in the ICUs of the Military Hospital (n=55)

| Variable | n | % | Variable | n | % |
| --- | --- | --- | --- | --- | --- |
| **Gender (n=55)** |  |  | **Renal function (n=55)** |  |  |
| Male | 28 | 50.9 | Normal | 33 | 60.0 |
| Female | 27 | 49.1 | Impaired | 22 | 40.0 |
| **Age in years (n=54)** |  |  | **Liver function (n=55)** |  |  |
| Median | 63.5 |  | Normal | 53 | 96.4 |
| Min-Max | 19-95 |  | Impaired | 2 | 3.6 |
| **Patients conditions (n=55)** |  |  | **Patients on vasopressors (n=55)** |  |  |
| Sepsis | 17 | 30.9 | No | 46 | 83.6 |
| Neurological | 8 | 14.5 | Yes | 9 | 16.4 |
| Cardiovascular | 8 | 14.5 | **Patients on steroids (n=55)** |  |  |
| Trauma | 8 | 14.5 | No | 43 | 78.2 |
| Stroke | 5 | 9.1 | Yes | 12 | 21.8 |
| Gastroenterology | 3 | 5.5 | **Patients on quinine I.V. (n=55)** |  |  |
| Cancer/Tumor | 2 | 3.6 | No | 54 | 98.2 |
| Endocrine | 2 | 3.6 | Yes | 1 | 1.8 |
| Respiratory | 2 | 3.6 | **On Fluoroquinolones (n=55)** |  |  |
| **Hyper glycaemia status (n=55)** |  |  | No | 51 | 92.7 |
| Non diabetic | 40 | 72.8 | Yes | 4 | 7.3 |
| Type 2 DM | 13 | 23.6 | **On atypical antipsychotics (n=55)** |  |  |
| Type 1 DM | 2 | 3.6 | No | 54 | 98.2 |
| **Feeding status (n=55)** |  |  | Yes | 1 | 1.8 |
| Enteral feeding | 32 | 58.2 |  |  |  |
| Oral feeding | 15 | 27.3 |  |  |  |
| Non per oral | 8 | 14.5 |  |  |  |

### Glycaemic status of ICU patients and hyperglycaemia control methods used in Military Hospital

A highly statistically significant association (Likelihood ratio = 49.964, *p* = 0.000) was found between the hyperglycaemia control method used and the diabetes status of the patients as indicated by table 5.

**Table 5:** Hyperglycaemia control methods by the glycaemic status of the patients (n=55)

| Method used | Hyperglycaemia status |  |  |  |  |  | Total |  | *p-value\** |
| --- | --- | --- | --- | --- | --- | --- | --- | --- | --- |
|  | Type 1 DM |  | Type 2 DM |  | Non-diabetic |  |  |  |  |
|  | n | % | n | % | n | % | n | % |  |
| Sliding scale | 0 | 0.0 | 9 | 81.8 | 2 | 18.2 | 11 | 20.0 | 0.000 |
| Basal-Bolus | 0 | 0.0 | 2 | 100.0 | 0 | 0.0 | 2 | 3.6 |  |
| Insulin I.V infusion | 0 | 0.0 | 1 | 100.0 | 0 | 0.0 | 1 | 1.8 |  |
| Other methods | 1 | 33.3 | 1 | 33.3 | 1 | 33.3 | 3 | 5.5 |  |
| None | 1 | 2.6 | 0 | 0.0 | 37 | 97.4 | 38 | 69.1 |  |
| **Total patients** | **2** | **3.6** | **13** | **23.6** | **40** | **72.7** | **55** | **100.0** |  |

\* Likelihood ratio=49.964

### Blood glucose levels and hyperglycaemia control methods used

Two classifications of random blood glucose (glycaemic levels and the NICE-SUGAR blood glucose levels) were used. The *glycaemic levels classification* revealed that 79.6% (43/54) of the patients were normal glycaemic (BG: 71-180 mg/dl), 18.5% (10/54) were hyperglycaemic (BG: > 180 mg/dl) and a patient (1.9%, 1/54) was hypoglycaemic (BG: < 71 mg/dl). In the other hand, the *NICE-SUGAR blood glucose classification* indicated that 61.1% (33/54) of the patients were below range (BG: < 140 mg/dl), 20.4% (11/54) were in within random glucose level (BG: 140-180 mg/dl), 18.5% (10/54) were above the range of BG > 180 mg/dl.

Regarding the hyperglycaemia control methods, they were used for 31.5% (17/54) of the patients. Table 6 revealed that 5.9% (1/17) of the patients was monitored in using the best appropriate method which was insulin infusion; 29.4% (5/17) of the patients were under alternative glycaemia control methods which were namely basal-bolus (11.8%, 2/17), mixed insulin (11.8%, 2/17) and Oral (5.9%, 1/17). Unfortunately, the majority of patients (64.7%, 11/17) had their glycaemia control based on the old fashion method of sliding scale despite a no statistically significant association (likelihood ratio = 10.108, *p* = 0.258) between the NICE-SUGAR targets and the method used.

**Table 6:**
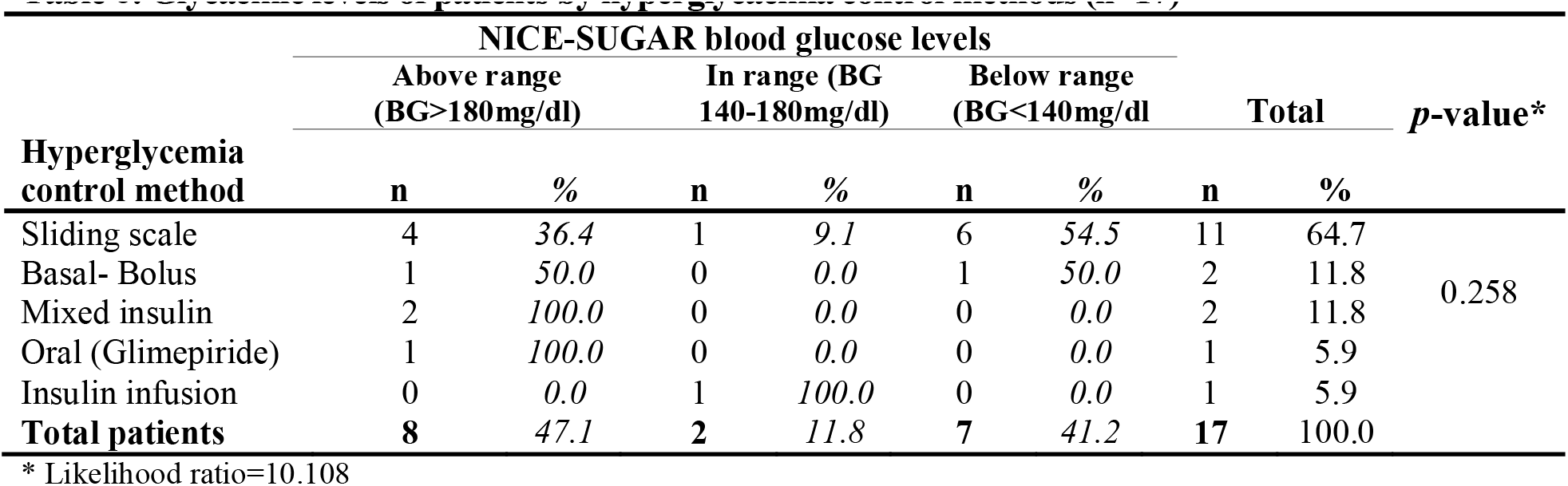
Glycaemic levels of patients by hyperglycaemia control methods (n=17)

## Discussion

More than half (66.7%, 14/21) of the doctors received a training on hyperglycaemia control, while, only 36.7% (22/60) of the nurses were trained with a statistically significant difference (*p* = 0.017) between the status of the staff and being trained on hyperglycaemia; training of health professionals is crucial to sustain evidence-based practice [30, 31].

In this study, regarding the knowledge of staff about hyperglycaemia control methods, there was no difference (*p* > 0.05) between the knowledge of doctors and nurses on Basal-Bolus and insulin infusion methods and their training status. This raised a question on the training programs implemented and emphasized the need for a standard updated policy with appropriate training material addressing the gaps of knowledge on hyperglycaemia control methods regardless the status of the staff [18].

Target blood glucose level of 140-180 mg/dl, acceptable for most ICU patients [11] and adopted by most of the major agencies [2, 3, 13] was known by only 11% of our study participants.

In our research, the practice of staff towards blood glucose monitoring frequency did not differ between trained and untrained doctors and nurses. This monitoring method using point of care (POC), glucometer or ICU laboratory, is acceptable as well as continuous glucose monitoring (CGM) [32]. However, regarding the practice towards DKA, as published in the literature [1, 19, 33], it statistically differed between doctors and nurses (*p* = 0.006), as well as, according to training status (*p* = 0.036).

Consistent with guideline-based practice [34, 35], all the staff (100.0%) of the cardiac care unit (CCU) in the Military Hospital were practicing the HbA1c measurement, as expected in such unit; contrary to the mixed (71.9%) and surgical ICUs (30.0%) with a statistically significant difference (*p* = 0.002) across the ICUs.

Detailed assessment on barriers and facilitators on policy implementation was published elsewhere [18, 31] as well as the availability of infusion pumps (indicated for the administration of insulin) in ICU [13, 34, 35]. Our findings revealed that the surgical ICU was the least equipped with an average of 1.7 infusion pumps±0.82.

The dominant hyperglycaemia control method in both surgical and mixed ICUs was sliding scale, which stood as the standard practice of our study participants while this method was discouraged [17, 19, 34, 36, 37]. Insulin infusion method is the recommended control method [1, 28, 22, 37], hence the need to move away nowadays from sliding scale [36]. This was our leitmotiv for proposing a protocol for glycaemia control in Sudan ICUs Military Hospital (supporting information S1). The proposed protocol is justified by our findings, which revealed that more than half of the care providers used sliding scale and were satisfied with it. Despite 89.0% of those caregivers did not know the target BG level [11] with a no statistically significant association (*p* = 0.365) between the staff satisfaction and the methods used. This appeal for the adoption of local institutional guidelines for all Military Hospital ICUs given the diversity of the specialities of health professionals [34].

Regarding patient safety, no glycaemia control method was used for the majority (69.1%, 38/55) of the patients; this was consistent with a Brazilian study [38] reporting the dominant use of sliding scale in ICUs; this reduced the use of basal-bolus and insulin infusion methods (69.2%, 7.1%, 3.9%).

The American Diabetes Association (ADA) [2] and Nottingham University Hospitals (NUH) guidelines [13], which pledged that hyperglycaemic patients even non-diabetics should have their glycaemia levels controlled, contradicted our findings and the ones of Moreira J.E.D et al. [38].

The blood glucose readings pointed that 11.8% of the patients had readings in the target range of 140-180 mg/dl, 41.2% had BG levels below the target range and 47.1% of the patients were hyperglycaemic (BG > 180 mg/dl). Our findings raised concerns about the nutritional status of the patients and the methods used as discussed in the literature [8, 9, 39]. In our research, insulin infusion method was used for one patient and the NICE-SUGAR target was achieved in-line with published data [6, 11]. While, mixed insulin method did not achieve the target glycaemic range as already reported by Marik P.E et al. [10]. Sliding scale method achieved the target range in only 9.1% of the patients of our study; consistent with published literature [8, 19, 40] recommending the use of insulin infusions in ICU patients to achieve the NICE-SUGAR range which had proven efficacy and safety in low-income countries [41].

The limitations of our study were due to the exclusion of the medical ICU because of renovation at the time of our data collection, and the data collected from working staff were not validated through Cronbach test of reliability. Nonetheless, the findings provided key elements that enabled the development of a protocol approved by the decision making ICU professionals and yet to be applied with respect to the forthcoming national guidelines.

## Conclusions

The poor knowledge and a lack of awareness towards hyperglycaemia management led to inappropriate implementation of glycaemia control methods across the Military Hospital ICUs. Sustained training programs on hyperglycaemia control for ICU healthcare staff are indicated as well as the availability of local guidelines on glycaemia control in all ICUs is needed. For best practice in ICUs, the use of intravenous insulin infusion targeting NICE-SUGAR blood glucose level should be recommended firstly, then, switching to basal-bolus method with nutritional support when patients get stable [6,39].

## Data Availability

To support data sharing and in compliance with MedRxiv data policy, all the data used in the framework of this research is deposit

## Declarations

### Ethical approval and consent to participate

SUMASRI Institutional Review Board of the University of Medical Sciences and Technology reviewed the proposal. Ethical Approval was obtained from the Military Hospital, the implementation of the research was granted by the administration of the respective ICUs. Participants (doctors, nurses and patients) were well informed about the research objectives and verbal consent was obtained from each of them. They were ensured about their confidentiality with the use of an anonymous research tool and that the data collected from them would be used strictly for the purpose of the study objectives.

### Consent for publication

Not applicable

### Availability of data and materials

To support data sharing and in compliance with PLOS data policy, all the data used in the framework of this research will be deposit to Dryad Digital Repository through to obtain a DOI which will be communicated to PLOS ONE.

### Authors’ contributions

#### AOHG

Elaborated the research proposal, implemented the field data collection, developed the local protocol for glycaemia control, conducted the statistical analysis and drafted the initial manuscript and the final manuscript.

#### AMAM

Facilitated the administrative arrangements at the Military Hospital, revised the proposal and read the final manuscript prior to submission.

#### NM

Supervised the implementation of the research from proposal to completion, and proof read the final the manuscript.

## Abbreviations

AACE: American Association of Clinical Endocrinologists
ACP: American College of Physicians
ADA: American Diabetes Association
BG: Blood glucose
CCR: Critical Care Room
CCU: Cardiac Care Unit
CGM: Continuous Glucose Monitoring
ICU: Intensive Care Unit
IIT: Intensive Insulin Therapy
NICE-SUGAR: The Normoglycaemia in Intensive Care Evaluation – Survival Using Glucose Algorithm Regulation
NUH: Nottingham University Hospitals
POC: point of care
SCCM: Society of Critical Care Medicine
SGC: Space Glucose Control

## Acknowledgments

The authors are grateful to the patients, doctors and nurses of the Military Hospital whose participation enabled this study to be completed.

## Supporting Information captions

### Document attached

**S1 File** Local guidelines for Hyperglycaemia Control in Intensive Care Units of Military Hospital, Sudan, 2020

### Tables

**S1 Table 1**: Characteristics of the healthcare staff and training on glycaemia control (n = 81)

**S2 Table 2**: Awareness of healthcare staff towards hyperglycaemia control methods and the reasons for lack of awareness (n = 81)

**S3 Table 3**: Management of diabetic ketoacidosis (DKA) by the participants according to their status of training on glycaemia control (n = 81)

**S4 Table 4**: Characteristics of patients admitted in the ICUs of Military Hospital (n = 55)

**S5 Table 5**: Hyperglycaemia control methods by the glycaemic status of the patients (n = 55) **S6 Table 6**: Glycaemic levels of patients by hyperglycaemia control methods (n = 17)

### Figure

**S1 Figure 1:** Hyperglycemia control methods used among healthcare professionals

